# Bioinformatics and system biology approach to identify the influences of COVID-19 on metabolic unhealthy obese patients

**DOI:** 10.1101/2023.01.25.23284996

**Authors:** Tengda Huang, Nan Jiang, Yujia Song, Hongyuan Pan, Jincheng Bai, Bingxuan Yu, Xiaoquan Li, Jinyi He, Kefei Yuan, Zhen Wang

**Affiliations:** Department of Liver Surgery & Liver Transplantation, State Key Laboratory of Biotherapy and Cancer Center, West China Hospital, Sichuan University, and Collaborative Innovation Center for Biotherapy, Chengdu, P.R. China

**Keywords:** SARS-COV-2, metabolic unhealthy obese, differentially expressed genes, gene ontology, protein-protein network (PPI), hub gene, drug molecule, gene-disease association

## Abstract

**Objective:** The severe acute respiratory syndrome coronavirus 2 (SARS-COV-2) has posed a significant challenge to individuals’ health. Increasing evidence shows that patients with metabolic unhealthy obesity (MUO) and COVID-19 have severer complications and higher mortality rate. However, the molecular mechanisms underlying the association between MUO and COVID-19 are poorly understood. We sought to implement transcriptomic analysis using bioinformatics and systems biology analysis approaches.

**Methods:** Here, two datasets (GSE196822 and GSE152991) were employed to extract differentially expressed genes (DEGs) to identify common hub genes, shared pathways and candidate drugs and construct a gene-disease network.

**Results:** Based on the identified 65 common DEGs, the results revealed hub genes and essential modules. Moreover, common associations between MUO and COVID-19 were found. Transcription factors (TFs)–genes interaction, and DEGs-miRNAs coregulatory network were identified. Furthermore, the gene-disease association were obtained and constructed.

**Conclusions:** The shared pathogenic pathways are noted worth paying attention to. Several genes are highlighted as critical targets for developing treatments for and investigating the complications of COVID-19 and MUO. Additionally, multiple genes are identified as promising biomarkers. We think this study’s result may help in selecting and inventing future treatments that can combat COVID-19 and MUO.

**Answer for the Study Importance Questions:** *What is already known about this subject?:* SARS-COV-2 infection can cause additional severe complications, particularly in patients with obesity and associated metabolic disturbance, which can also increase the risk of SARS-COV-2 infection and hospitalization. SARS-COV-2 infection can cause additional severe complications, particularly in patients with obesity and associated metabolic disturbances, which can also increase the risk of SARS-COV-2 infection and hospitalization.

*What are the new findings in your manuscript?:* Based on the 65 identified common DEGs, the shared pathogenic pathways are noted worth paying attention to. Several genes are highlighted as critical targets for developing treatments for and investigating the complications of COVID-19 and MUO. Additionally, multiple genes are identified as promising biomarkers. We think this study’s result may help in selecting candidate drugs and inventing future treatments that can combat COVID-19 and MUO.

*How might your results change the direction of research or the focus of clinical practice?:* Potential pathways and genes that significantly affect the prognosis of COVID-19 patients with MUO were identified, which might be helpful for further research about the detailed mechanism of how obesity affects the coronavirus infection. Additionally, the extracted candidate drugs might be the potential drugs for treating these two diseases in clinical practice. The gene-disease network also revealed essential genes linking them with other diseases, providing information for complications studies.

## Introduction

Severe acute respiratory syndrome coronavirus 2 (SARS-COV-2) is a highly contagious coronavirus responsible for the life-threatening COVID-19. By December 2022, more than 650 million people were infected by SARS-COV-2, and it also caused more than 6 million deaths in more than 300 countries and regions around the world[1]. The risk of severe symptoms and complications, mortality and hospitalization in COVID-19 patients have been recently reported to be significantly increased due to some unhealthy pre-existing conditions, including hypertension, type 2 diabetes, and obesity[2]. Compared with metabolic healthy patients, patients with metabolic disturbances also were observed to have a much poorer prognosis[3]. Additionally, it has been reported that obesity can severely hamper the immune cells responsiveness to weaken the long-term protection against SARS-COV-2[4]. Metabolic disturbances are a common complication of obesity, and only 15% to 20% of obese patients are not affected by them[3]. Metabolic disturbances have been proven to be a significant aggravating factor in many obesity-related diseases, so the underlying mechanisms linking COVID-19 and metabolic unhealthy obesity (MUO) are needed to be better clarified.

Angiotensin Converting Enzyme 2 (ACE2), whose gene expression is founded in many human tissues, has been known to play a vital role as a receptor for SARS-CoV-2 entry and infection in target cells[5]. MUO is a systemic disease, which has been discovered to be associated with the upregulated ACE2 expression in tissues in the whole human body[3]. As reviewed by Goossens et al., with the increased number of adipose tissues, the ACE2 expression is upregulated, making the concentration of SARS-COV-2 in adipocytes significantly higher than in other tissue cells, becoming a viral reservoir for SARS-CoV-2[6]. Additionally, the contribution of MUO related upregulated Transmembrane Serine Protease 2 (TMPRSS2) expression, hyperglycemia, and weakened immune surveillance to the poorer prognosis of COVID-19 in the corresponding patient population are also being in-depth investigated[3]. This serves to demonstrate the potential importance of interaction between MUO and COVID-19.

In this study, we investigated the interaction of COVID-19 with MUO. Two datasets were utilized to discover it. Those datasets were collected from the Gene Expression Omnibus (GEO) database, where GSE196822 for COVID-19 and GSE152991 for MUO. Shared Differentially Expressed Genes (DEGs) were identified through cross-analysis of the two datasets, and the result was used to discover the relevant signaling pathways as well as the potential genes that are therapeutic targets for patients with MUO and COVID-19. Also, efforts to reveal the molecular mechanisms linking MUO and COVID-19 were made, including predictive transcription factor (TFs)-gene interaction, protein-drug interaction, and DEG-miRNA network analysis based on the identified DEGs. We further determined the association with other diseases that might provide further insights for the study about complications of and treatments for COVID-19. We hope that the findings of this study will provide preliminary information that may help to understand the interaction between COVID-19 and MUO and help in selecting proper drugs and inventing future treatments that can combat COVID-19 and MUO.

## Methods

### Gene expression datasets

In this study, the RNA-seq data were obtained from the GEO database (https://www.ncbi.nlm.nih.gov/geo/) of the National Center for Biotechnology Information (NCBI). The GEO accession ID of the COVID-19 dataset is GSE196822, which is transcriptional profiling of COVID-19 whole blood samples through high throughput sequencing Illumina HiSeq 4000 (Homo sapiens) for RNA sequence extraction. The COVID-19 dataset contains thirty-four infection samples and nine healthy controls. Besides, the GSE152991 dataset consists of twenty-two metabolic unhealthy obese subcutaneous adipose samples and eleven healthy controls. The adipose samples were sequenced by a high-throughput sequencing system called Illumina NovaSeq 6000 (Homo sapiens).

### Identification of DEGs and common DEGs among COVID-19 and MUO

A gene is characterized as DEG when tested at the transcriptional level under different conditions, and there were significant differences. The purpose of this analysis is to obtain DEGs for the datasets. The DEGs from GSE196822 were identified using DESeq2 of R programming language[7]. In the analysis of the MUO dataset, the limma package[8] was used to determine differentially DEGs between the metabolically healthy lean group (MHL) and the MUO group. Cutoff criteria (False Discovery Rate (FDR) < 0.05 and |log_2_ Fold Change| ≥ 1 was applied to obtain significant DEGs from both datasets. VENN analysis was performed using an online analysis tool called Jvenn (http://bioinfo.genotoul.fr/jvenn.) to acquire the common DEGs of GSE196822 and GSE152991.

### Gene ontology and pathway enrichment analysis

EnrichR (https://maayanlab.cloud/Enrichr/) was utilized to conduct gene ontology (biological process, cellular component and molecular function) and pathway enrichment analysis of common DEGs. Pathway enrichment analysis concluding Kyoto Encyclopedia of Genes and Genomes (KEGG), WikiPathways, Reactome and BioCarta were performed to identify the overlapped pathways among MUO and COVID-19. The *P*-value < 0.05 was taken as a metric for quantifying the top-listed pathways and gene ontological processes.

### Protein–protein interactions analysis

The common DEGs were used to construct protein subnetworks and further reveal the protein interactions between MUO and COVID-19. STRING—a protein interactome database is used in this analysis. The protein–protein interaction (PPI) network was constructed and visually represented by Cytoscape (v3.9.1).

### Hub gene extraction and submodule analysis

Nodes, edges, and their connections compromise the PPI network. The nodes are the essential component of the network, and the most involved ones are regarded as the hub genes. Cytohubba (http://apps.cytoscape.org/apps/cytohubba)[9] is a novel Cytoscape—plugin for ranking and extracting potential or targeted elements of a biological network based on various network features. Among the 11 methods provided by Cytohubba that can be used to investigate networks from viewpoints, Maximal Clique Centrality (MCC) has the best accuracy and effectiveness, which is used to identify the top hub genes from the PPI network.

### Recognition of transcription factors (TFs) and miRNAs engaged with common DEGs

The transcription rate of a gene is closely related to the regulation of the TFs that attach to it. Therefore, TFs are very important for exploring gene activity at the molecular level. The topologically credible TFs are located using the NetworkAnalyst platform (3.0) (http://www.networkanalyst.ca) from the JASPAR database (http://jaspar.genereg.net/). Further, miRNAs are the nucleotides that attach to gene transcripts to regulate protein expression. The miRNAs–gene interaction networks were constructed via NetworkAnalyst and illustrated in Cytoscape. The topological analysis of miRNAs was performed to specify the most influential miRNAs from mirTarbase.

### Gene–disease association analysis

DisGeNET is a comprehensive database of gene–disease associations that synchronizes the biomedical characteristics of many diseases to determine the relationship between genes and specific diseases. It emphasizes the critical role of key genes in the occurrence and development of diseases. The gene-disease relationship via NetworkAnalyst was examined to reveal correlated diseases and their complications with hub genes based on the DiGeNET database.

## Results

### Identification of DEGs and common DEGs among MUO and COVID-19

To study the relationship and interaction between MUO and COVID-19, an analysis of the human transcriptomic datasets from the GEO database was conducted to identify and classify the pathogenic genes that stimulate MUO and COVID-19. Table 1 lists the summarized information of the identified DEGs from the datasets. Firstly, 1,668 DEGs were identified for COVID-19, including 839 up-regulated and 829 down-regulated DEGs. In the same way, 441 up-regulated and 108 down-regulated DEGs for MUO were selected after completing the different statistical analysis processes. Secondly, 65 common DEGs from MUO and COVID-19 datasets were identified after performing the Venn analysis. The result suggested a certain correlation between the two diseases as they share several common genes.

**Table 1.**
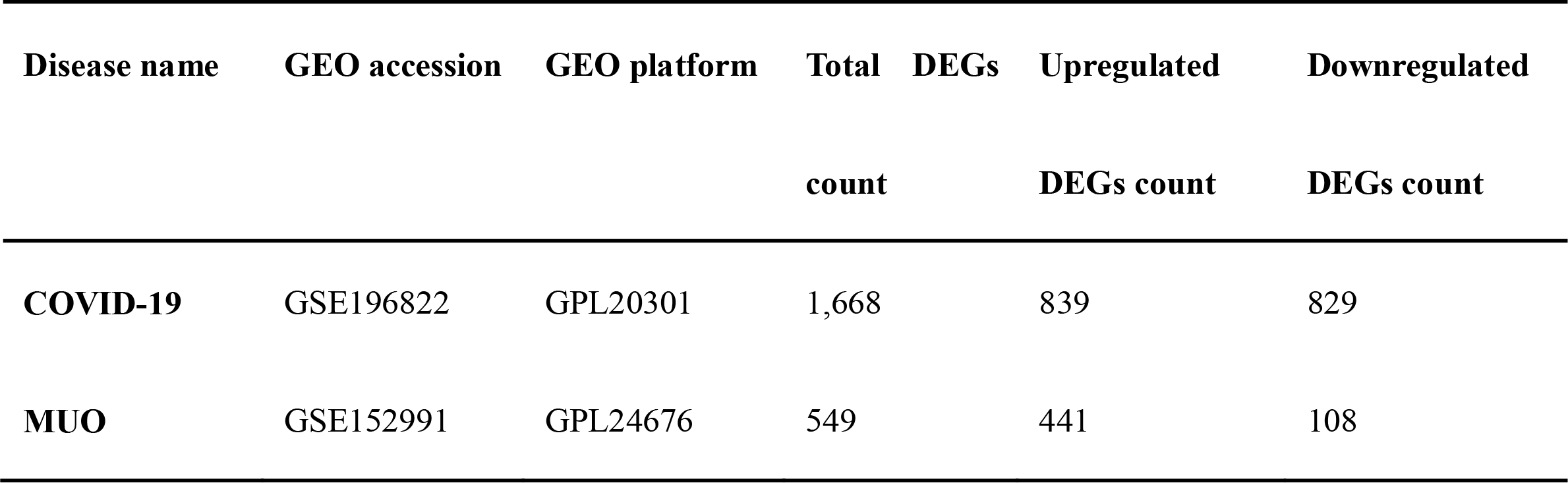
Overview of datasets with their geo-features and their quantitative measurements in this analysis

### Gene ontology and pathway enrichment analysis

To further investigate the functions and components of common DEGs, the Gene ontology analysis has been performed. The top 10 terms in the biological process, molecular function and cellular component categories are summarized in Table 2. The bar graphs for top ontological terms in each category are displayed linearly in Figure 1(A-C).

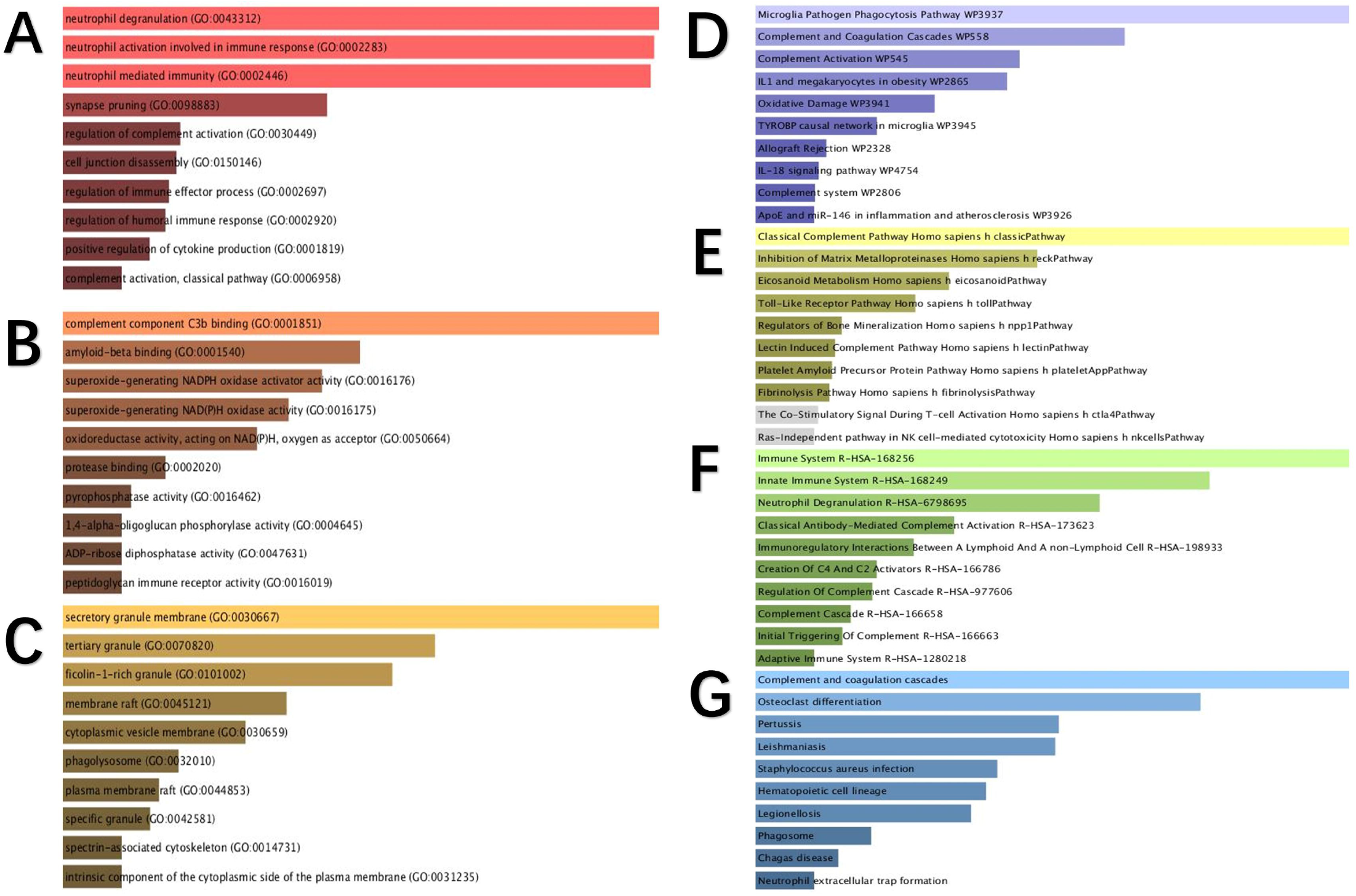

**Table 2.**
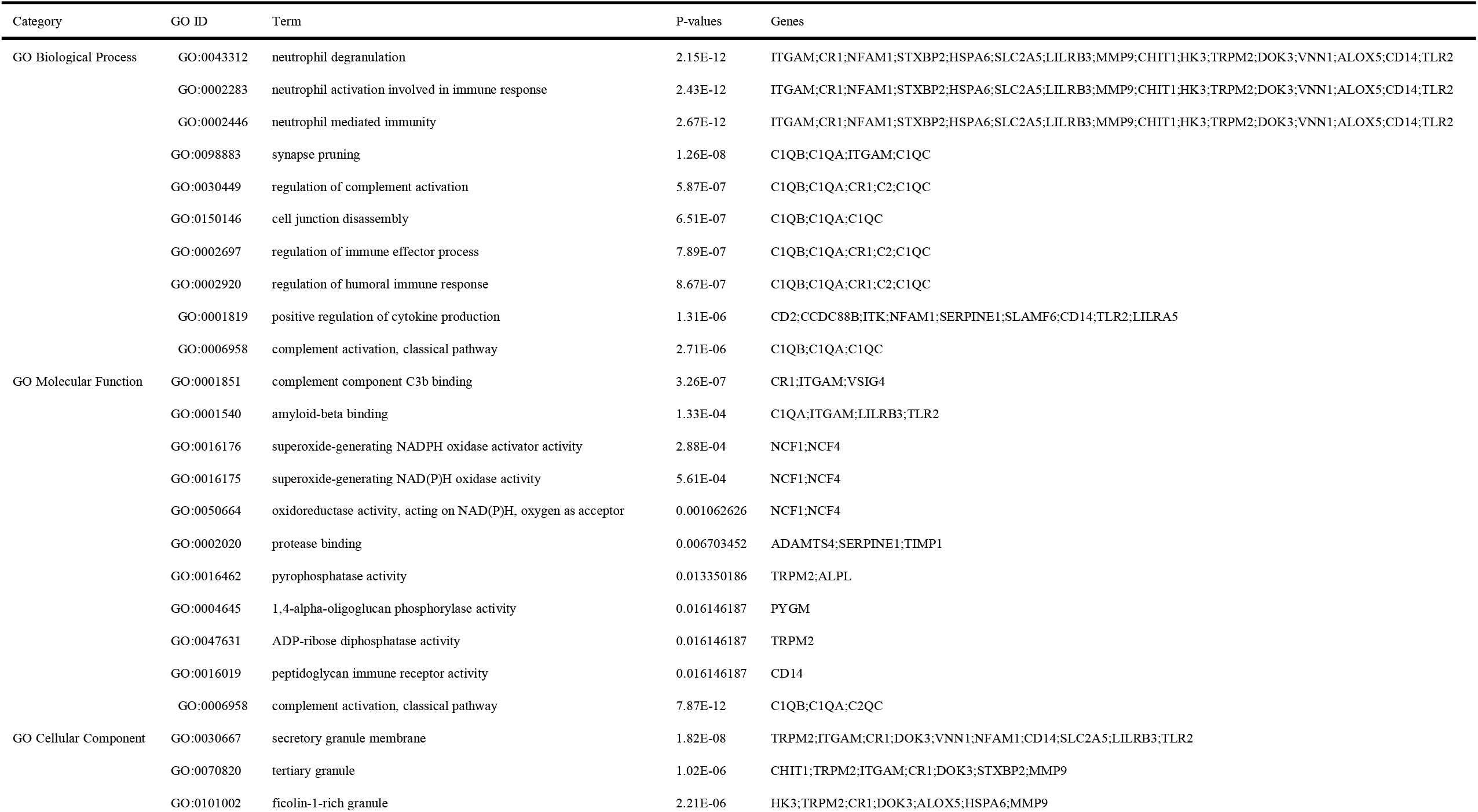
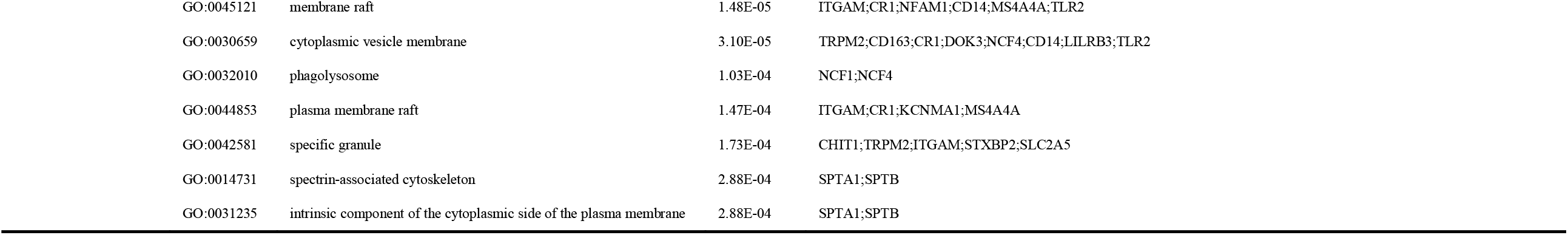
Ontological analysis of common DEGs between MUO and COVID-19.

In addition, pathways analysis was performed to reveal the interaction between MUO and COVID-19 through basic molecular or biological processes. The top 10 enriched pathways of the shared DEGs obtained from the selected database are enlisted in Table 3. The bar graphs of the characterized pathway analysis for each database are displayed in Figure 1(D~G).

**Table 3.**
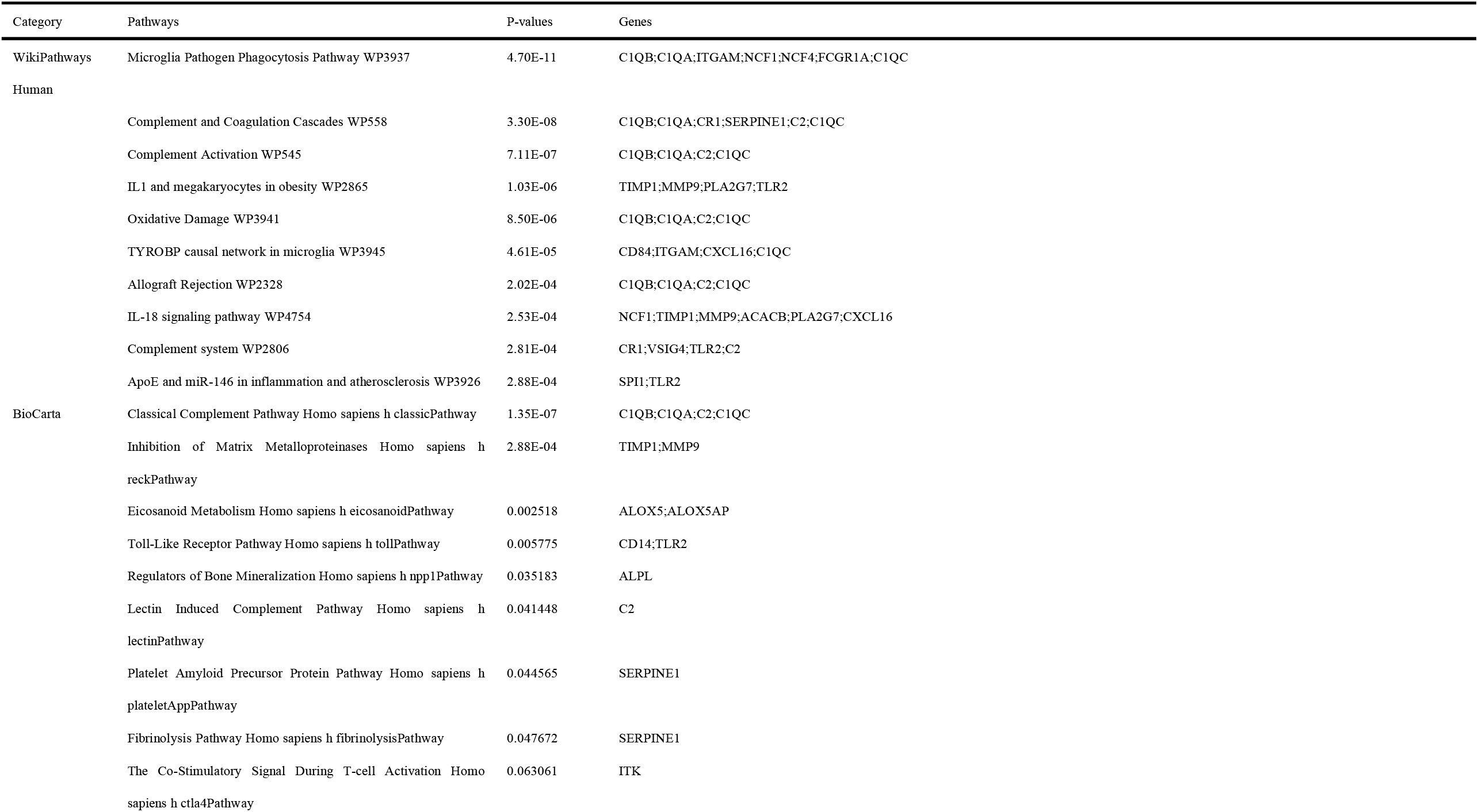
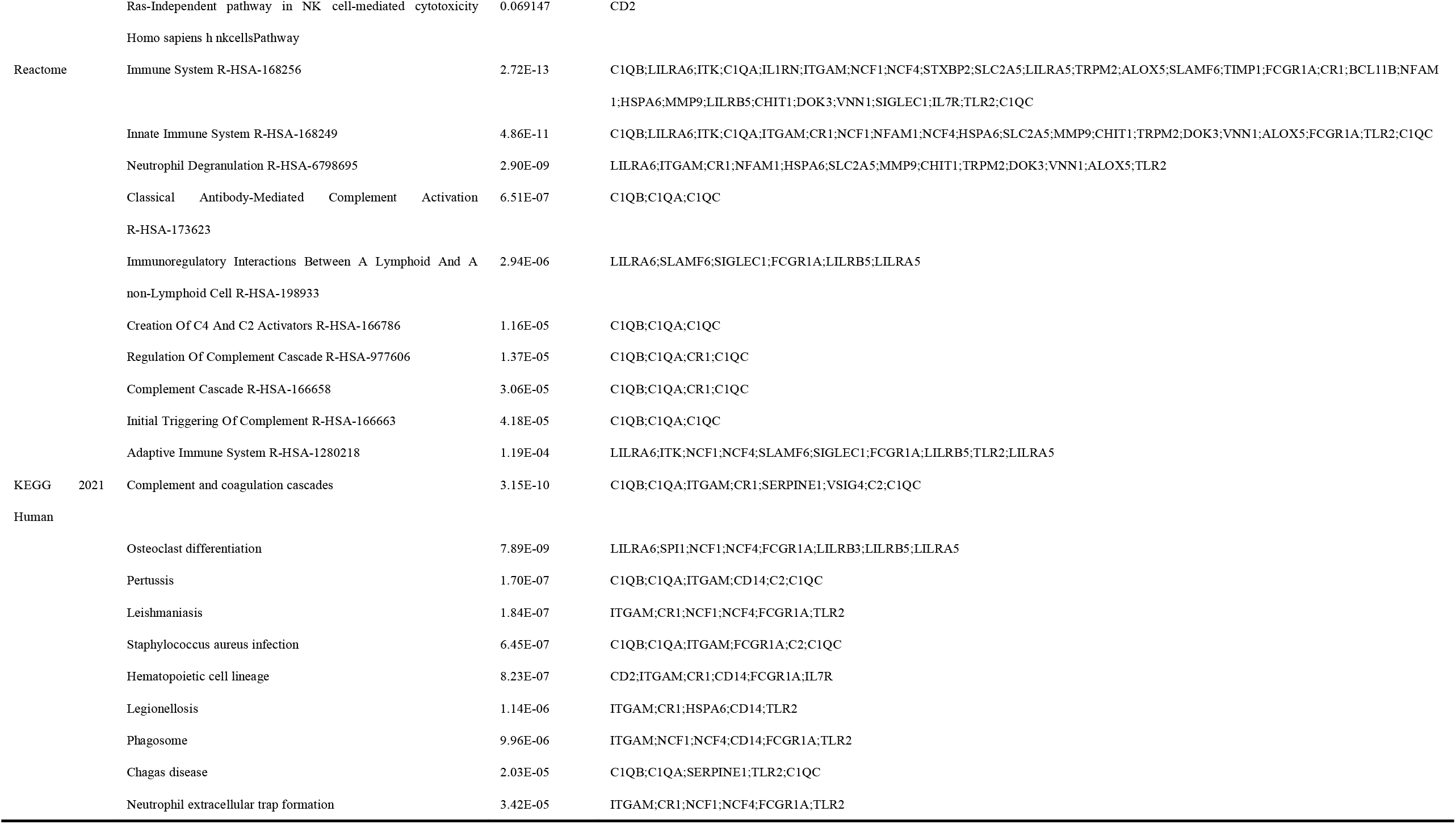
pathway enrichment analysis of common DEGs between MUO and COVID-19.

As seen in Table 2 and Figure 1(A~C), the most significant ontology pathways identified in the interaction between COVID-19 and MUO are primarily correlated with immune system response, including complement and microglial phagocytosis pathways.

### Classification of hub genes and submodules

To study the mechanisms of the interactions and connectivity between DEGs, the PPI network was constructed. This PPI network consists of 65 nodes and 167 edges and all the interconnected nodes are depicted in Figure 2(A). The most interconnected nodes in the PPI network are acknowledged as hub genes. The top 10 identified hub genes are namely SPI1, CD163, C1QB, SIGLEC1, C1QA, ITGAM, CD14, FCGR1A, VSIG4 and C1QC. These hub genes can be the potential biomarkers and therapeutic targets, so a submodule network was constructed to understand their near connectivity and proximity deeper. cc

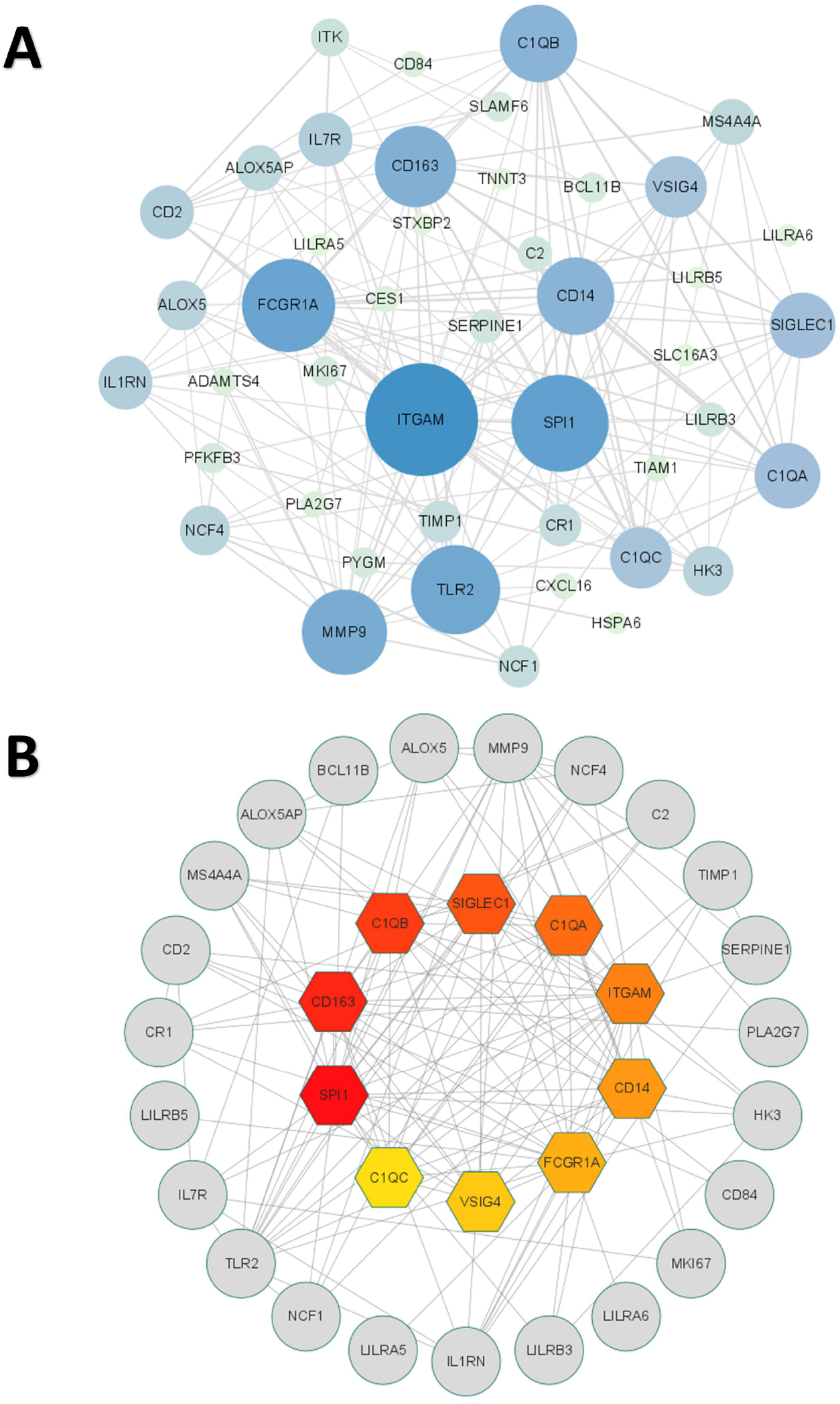

### Identification of DEGs–miRNA and TF–gene interactions for the common genes

To understand the interaction between COVID-19 and MUO at the transcriptional level and further investigate the transcriptional regulatory molecule, the regulator TFs and miRNAs were decoded through a network-based approach. The interaction network between common DEGs and TFs is depicted in Figure 3(A). cc From the interaction network analysis, 65 TFs and 47 miRNAs were found to be the regulatory signals that regulate multiple identified hub genes, implying that the transcriptional regulators play an essential role in the interaction between MUO and OCIVD-19.

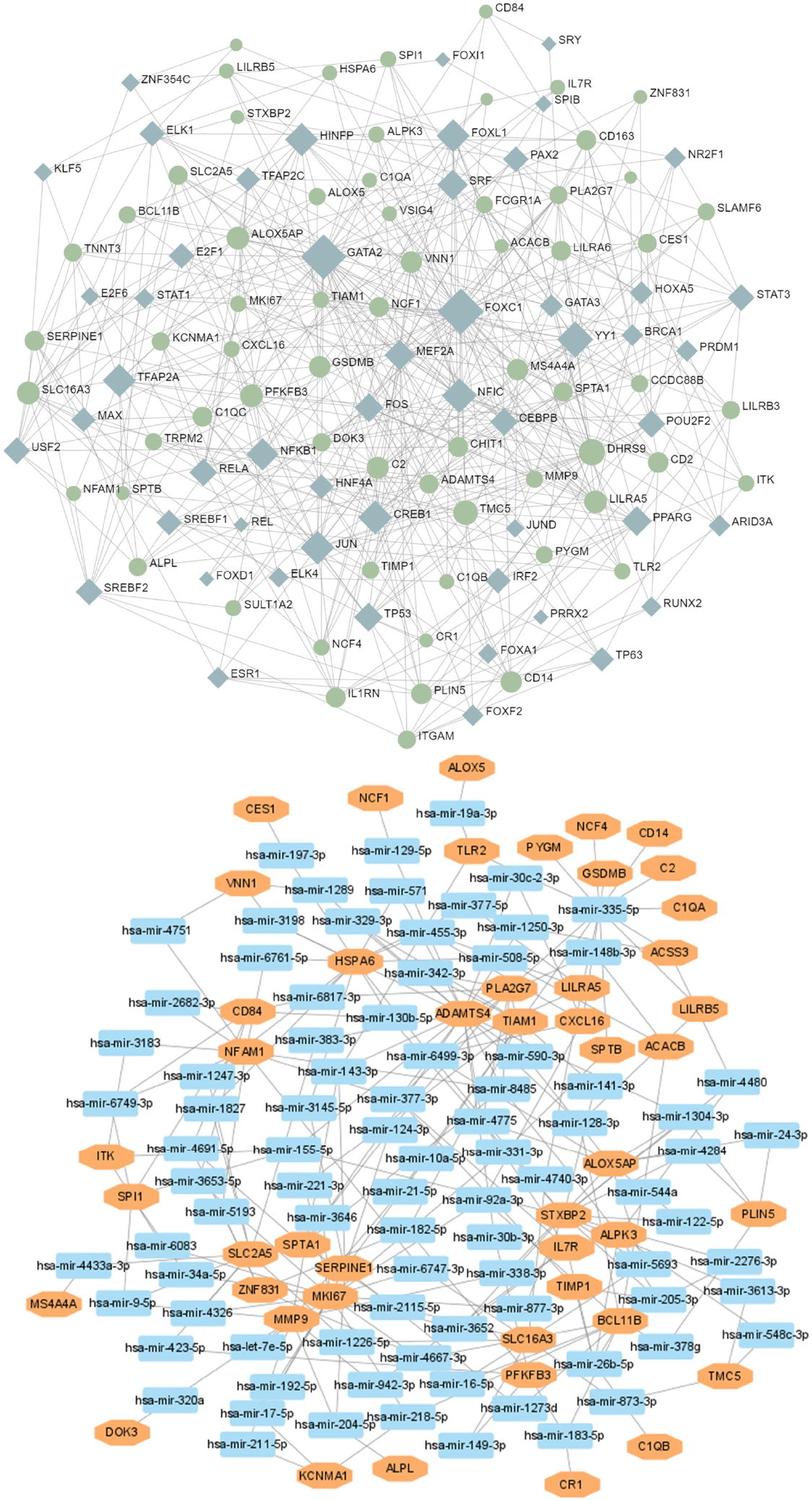

### Identification of disease associations

Different diseases can be associated when their overlapped pathogenic gene. Additionally, the disease treatments are designed based on the relationship between genes and diseases. In order to further identify the diseases most related to MUO and COVID-19, the gene-disease relationship analysis was conducted to review the disease that were most coordinated with our reported hub genes. It showed that bipolar disorder, systemic lupus erythematosus, schizophrenia, and liver cirrhosis were most coordinated with the hub genes identified in this study. The multiple gene-disease relationships are depicted in Figure 4.

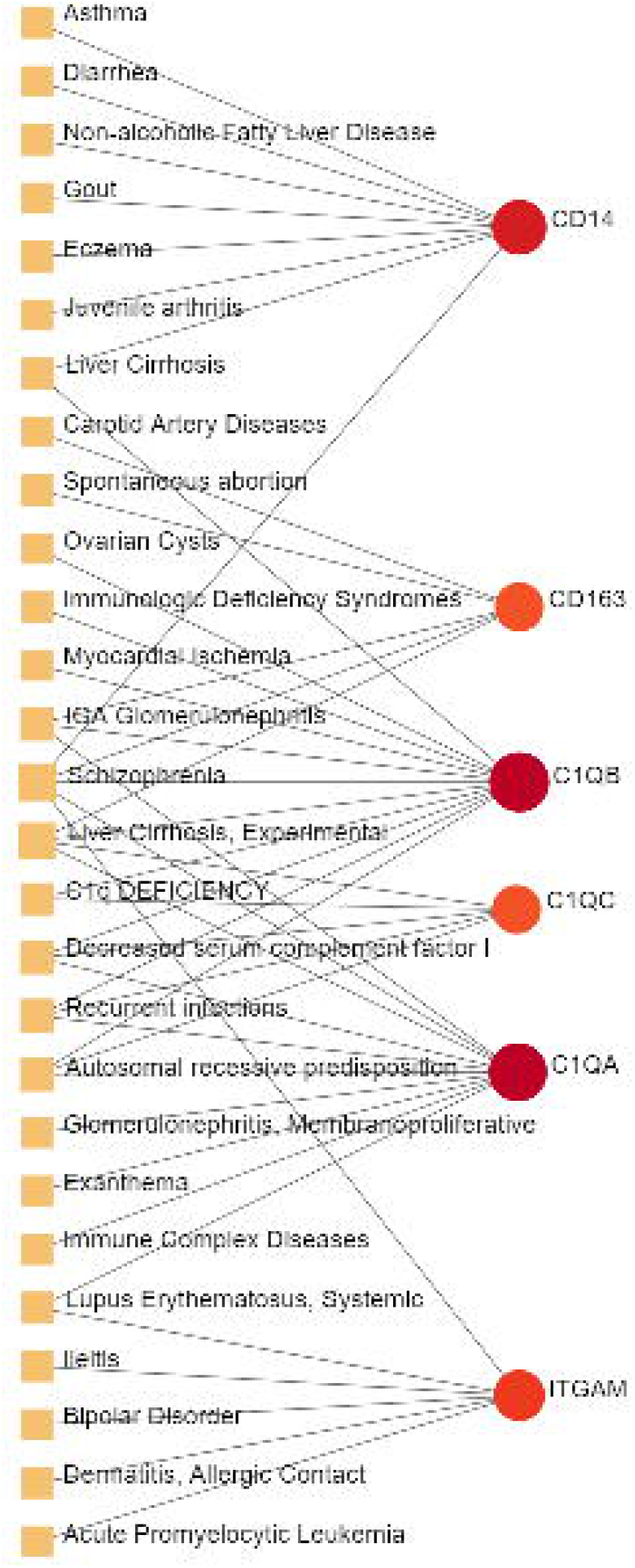

## Discussion

MUO is a type of metabolic disorder regarded as the main obesity-related complication. Patients with MUO usually preserve insulin resistance, hypertension, chronic inflammation, and altered liver inflammation[10], all of which are negative preconditions for the prognosis of COVID-19 infections[3].

Here, MUO and COVID-19 transcriptomics analysis revealed that 65 shared DEGs show similar expression patterns in these two disorders. GO pathway analysis was conducted to assess the biological importance of the identified common DEGs in the pathogenesis of MUO and COVID-19. Three types of GO analysis were conducted with the selected database as an annotation source in the ontological processes.

Neutrophil-related pathways are among the top GO terms for the biological process, including neutrophil degranulation and activation. Increasing evidence suggests that inhibiting neutrophil degranulation is beneficial for ameliorating inflammation-induced myocyte damage, hepatic acute phase response and thrombosis formation in severe COVID-19 patients[11]. In addition, a case study has shown that the upregulation of genes involved in neutrophil degranulation is the main reason for the side effect following vaccination[12].

In the molecular function experiment, complement component C3b binding (3 genes) and amyloid-beta binding (4 genes) are two top GO pathways. Concerning COVID-19, C3b recruits immune cells to the sites of infection and induces activation and further differentiation towards an inflammatory phenotype with the subsequent activation of lectin pathway (LP)-mediated C3b deposition, which is critical for the induction and maintenance of a severe inflammatory response to SARS-CoV-2[13]. Moreover, a previous study found that the raised level of C3b in serum is linked to obesity[14], which may provide essential insight into the severe inflammation that is occurred in patients with both MUO and CPVID-19. In addition, recent research has proved that SARS-CoV-2 infection elevates neuroinflammation by inducing dysregulation of microglia and astrocyte subpopulations, alters brain structure leads to abnormal accumulation of amyloid-beta[15]. Similarly, the pathway of activating and modifying proinflammatory cells in the central neuron system (CNS) is also the way that MUO increases the risk of AD[16]. It implicated the underlying synergistic effect of increasing AD occurrence on COVID-19 patients with MUO.

The top GO terms based on the cellular component were “secretory granule membrane” (10 genes) and “tertiary granule” (7 genes). Besides the neutrophil degranulation, which has been discussed above, the secretory granule is essential for SARS-COV-2 exiting the cell. Intracellular transport of SARS-COV-2 within the cells involves secretory granules, each containing a single virus particle. These vesicles then fuse with the plasma membrane of host cells, releasing the virus outside the cell and facilitating the transmission of SARS-COV-2[17]. Yeming Yang et al., revealed that metabolic unhealthy disorders could alter the membrane phospholipid distribution in multiple physiological systems[18], affecting secretory granule membrane synthesis.

The top pathways terms from the four selected databases were complement and coagulation cascades, osteoclast differentiation, microglia pathogen phagocytosis and inhibition of Matrix Metalloproteinases (MM). Here, complement and coagulation cascade activation are the common pathways in virus infection and metabolic disorders. COVID-19 induces the pro-inflammatory state, which stimulates uncontrolled activation of the complement system and neutrophil extracellular traps (NETs)-formation, both of which promote the coagulation cascade and induce a state of “thrombo-inflammation”[19]. In patients with MUO, the dysregulation of secreted proteins and the secretory machinery in the liver lead to the abnormally regulated complement and coagulation cascades[20], suggesting that this cascade can be an important target for developing alternative management measures for patients with both MUO and COVID-19. In addition, recent studies about patients diagnosed with COVID-19 have reported bone loss. Preliminary research showed that it might be due to the SARS-COV-2-induced cytokine dysregulation, as the circulating pro-inflammatory cytokines upregulate osteoclast differentiation in bone tissues[21]. As it is, it can aggravate lipid-altering conditions related to bone loss in MUO patients[22].Based on the identified DEGs, a PPI network was constructed to analyze interconnected proteins’ functional characteristics in-depth and predict potential drug targets. The hub genes essential in the pathogenesis of COVID-19 and MUO could be key drug targets and biomarkers in them. The top 10 hub genes associated with COVID-10 and MUO were retrieved through MCC method, including SPI1, CD163, C1QB, SIGLEC1, C1QA, ITGAM, CD14, FCGR1A, VSIG4, C1QC. Protein SPI1 and GATA2 are the essential transcription factors used to predict dysregulated hematopoiesis in bone marrow in severe COVID-19 patients. Recent evidence has proved the link between hematopoietic dysfunction of bone marrow and COVID-19, which is characterized by the accumulation of immature myeloid progenitors and a dramatic reduction of lymphoid progenitors[23]. Obesity-related abnormal glucose metabolism has also been reported to impair bone marrow function, especially hematopoiesis[24]. Therefore, patients with MUO precondition are more likely to suffer from severe SARS-COV-2 infection complication.

Another hub gene, which encoded CD163, indicates the importance of macrophage mediated SARS-COV-2 related inflammation as one of the pathogeneses of inflammation in the respiratory tract[25]. Daniel Wendisch et al., described the CD163-expressing macrophage phenotype as the profibrotic transcriptional phenotype during COVID-19 infection[26], which is also the reason why obese patients develop metabolic diseases[27]. Furthermore, the hub genes C1QA, C1QB, C1QC, VSIG4 and CD14 have been regarded as the main biomarkers for predicting macrophage and neutrophil mediated inflammation, especially in the respiratory and cardiovascular systems[28]. They are also the essential biomarkers that have been identified in the unfavorable metabolic profile of MUO patients[29]. Additionally, during the recovery process of patients, especially in the early recovery stage (ERS), the ratio of classical CD14^++^ monocytes was reported to be elevated, accompanied by high expression of inflammatory genes, which prevented patients from achieving a better prognosis. The hub protein ITGAM is an essential molecular receptor that plays a vital role in many complement mediated pathways that aggravate the COVID-19 symptoms, such as the C5a-C5aR1 axis in the pathophysiology of acute respiratory distress syndrome[30]. It is also a critical marker for inflammatory macrophages, whose infiltration can be elevated due to obesity[27]. Bronchoalveolar lavage fluid CD4+ T cells of patients with COVID-19 were T_H_1-skewed and showed de-repression of genes downregulated by VitD, which notably activates the recruiting of c-JUN and switches on the pro-inflammatory programs of T_H_1 cells[31]. This c-JUN mediated cascade has also been discovered to be activated by increased metabolic stress caused by obesity[32]. Based on this information, we can speculate that the induced raised degree of macrophage infiltration and transcriptional factors activation by glucose metabolism and oxidative phosphorylation can lead to a poorer prognosis and higher mortality of COVID-19.

Moreover, an LP that enhances ACE2-dependent infection by SARS-CoV-2 and induces neutralization by different classes of spike-specific antibodies has been identified in macrophages expressed the hub protein SIGLEC1[33]. Furthermore, cytokines storm is the main risk factor that leads to the death outcome of COVID-19 patients. CASP1 is the most involved pathway in cytokine storm, and YY1 is a crucial TF in CASP1 expression[34]. In obese mouse models, YY1 was markedly upregulated[35], which may provide us with the underlying cause of increased mortality in COVID-19 patients with MUO.

The common DEGs have also been identified by analyzing the TFs–gene and DEGs-miRNAs interaction. The result of this work, we have identified TFs, such as DHRS9, NFIC, CREB1, HINFP and FOXL1, which are highly expressed in the association between COVID-19 and MUO. Regarding determining the relationship between DEGs and miRNAs, hsa-mir-335-5p, hsa-mir-26b-5p, and hsa-mir-8485, have been identified among some of the miRNAs shared COVID-19 and MUO. Although the miRNA features of different COVID-19 complications have been determined using the systems biology approach, more studies still need to be conducted to investigate the effect of specific miRNA on SARS-COV-2 infection.

The gene-disease (GD) analysis was performed to predict relationships between common DEGs and different disorders. The analysis showed various diseases correlated with MUO and COVID-19, including mental illnesses, gastrointestinal tract, and immune and cardiovascular system disorders. For example, several genes, which are mainly related to schizophrenia and bipolar disorder, associated with mental disorders were founded in the study. The mortality rate of patients with mental disorders complicated with COVID-19 increased significantly, among which schizophrenia and bipolar disorder were the two highest risk factors[36]. Additionally, schizophrenia and bipolar disorder are also high-risk factors in MUO, because they can affect genetic and neuroendocrine processes to induce more weight gain[37]. Liver cirrhosis has also been founded in our GD network; the patients with it have a higher probability of severe COVID-19 symptom, particularly high rates of hepatic decompensation and death[38]. Furthermore, COVID-19 infection and MUO have been shown to cause and aggravate various chronic liver diseases[39]. Moreover, the involvement of myocardial ischemia in COVID-19 is directly associated with cytokine-mediated plaque destabilization and hypercoagulability, which induce ischemic stroke and acute myocardial infarction[40]. Additionally, there is a significant correlation between metabolic disorders and prolonged QTc interval[41], which is the early marker of transient myocardial ischemia.

In addition to presenting some interesting findings, our study has some limitations. These results, including DEGs and candidate drug identification, as well as all the network analysis, were obtained by bioinformatics and system biology approach. The results have not been verified using any animal model or human sample. Additionally, the selected datasets include different groups of people with two different diseases, rather than the same population with both MUO and COVID-19. Thus, further studies and clinical trials are needed to validate the biological functions of the identified hub genes and the safety and efficacy of the specified candidate drugs, as well as their pharmacological characteristics.

## Conclusions

In conclusion, for further investigation into the association between MUO and COVID-19, neutrophil and complement mediated pathways should be paid attention to. GATA2, YY1, JUN, VISG4, and SPI1 are highlighted as the important targets for developing treatment for patients with MUO and COVID-19 and investigating the several complications of COVID-19 and MUO, including Schizophrenia, bipolar disorder, chronic liver diseases and IgA glomerulonephritis. Additionally, CD14, CD163, ITGAM, C1QA, C1QB, C1QC and VSIG4 are identified as promising biomarkers for predicting the prognosis of COVID-19 and MUO.

## Supporting information

STREGA checklist

## Data Availability

All data produced in the present study are available upon reasonable request to the authors

