## Supplemental Table1 for "Bioinformatics and system biology approach to identify the influences of COVID-19 on metabolic unhealthy obese patients"

| **NAME OF DATABASE** | **REFERENCE** |
| --- | --- |
| GEO database | [1] |
| KEGG | [2] |
| WiKiPathways | [3] |
| Reactome | [2] |
| BioCarta | [4] |
| STRING | [5] |
| JASPAR | [6] |
| miTarbase | [7] |
| DsigDB | [8] |
| DrugBank | [9] |
| Comparative Toxicogenomics Database | [10] |
| DisGeNET | [11] |
